# At-home ankle and wrist accelerometry and machine learning capture motor impairment and disease progression in ataxia-telangiectasia

**DOI:** 10.64898/2026.11.21.26368661

**Authors:** Divya D. Kulkarni, Annika Ruth Toivonen, Anna Luddy, Sara Reiling, Jennifer Thornton, Franziska Hoche, Anoopum S. Gupta

**Author notes:** Correspondence to: Dr Anoopum S. Gupta, Department of Neurology, Massachusetts General Hospital, Boston, MA, USA.

## Abstract

**Background:** Ataxia-telangiectasia (A-T) is a rare, childhood-onset neurodegenerative disorder for which sensitive, age-appropriate motor outcome measures are needed to support emerging clinical trials. Clinician-rated ataxia scales are limited by subjectivity, low granularity, and floor and ceiling effects. We evaluated whether at-home wearable accelerometry, combined with machine-learned composite scores derived from submovement features, captures cross-sectional motor impairment and longitudinal disease progression in A-T.

**Methods:** 43 children with classic A-T (median age 9.0 years; IQR 5.7-13.4) and 50 age- and sex-matched controls wore GENEActiv accelerometers on the ankle and wrist during one week of natural behavior at home, repeated at 6- to 12-month intervals. From 85 submovement, activity-intensity, and spectral features per sensor site, we trained composite severity models targeting the Brief Ataxia Rating Scale (BARS) gait subscore (ankle) and BARS total score (wrist).

**Findings:** Ankle composite scores correlated strongly with BARS gait (r = 0.88), BARS total (r = 0.88), and caregiver-reported CPCHILD (r = -0.69); wrist composite scores correlated with BARS total (r = 0.88), BARS arm (r = 0.84), and CPCHILD (r = -0.78). Both models discriminated A-T from controls with large effect sizes (ankle Cohen’s d = 2.28; wrist d = 2.98) and demonstrated excellent within-week reliability (ICC = 0.95 ankle; 0.92 wrist). The ankle composite captured disease progression sensitively (Mean-to-Standard Deviation Ratio [MSDR] = 0.98; p < 0.0001). In a matched subset with both sensor and clinical data, ankle composite slopes significantly captured progression (Wilcoxon vs zero, p = 0.0036), whereas BARS total (p = 0.079) and BARS gait (p = 0.30) did not.

**Interpretation:** At-home ankle-based wearable accelerometry provides a sensitive, interpretable, and ecologically valid measure of motor function and disease progression in A-T and may serve as a candidate outcome measure for clinical trials.

**Research in context:** *Evidence before this study:* We searched PubMed and Google Scholar for articles published up to September 2026 using the terms "ataxia-telangiectasia", "wearable", "accelerometer", "digital biomarker", and "motor outcome measure". Clinician-rated ataxia scales remain the standard for assessing motor severity in A-T, but are limited by rater subjectivity, low granularity, and age-dependent baseline scores in children. Task-based wearable assessments have shown promise but share the limitations of requiring structured protocols and participant cooperation. At-home wearable accelerometry using submovement analysis had been applied to the wrist in children with A-T in one prior study by our group, demonstrating cross-sectional validity and longitudinal change over one year. Ankle-worn sensors had been used to estimate ataxia severity in children with A-T during a supervised gait assessment, and at-home ankle-based submovement measures and machine-learned composite scores had shown strong sensitivity in adult cerebellar ataxias. However, at-home composite scores and ankle-based measures had not been evaluated in A-T, and no study had compared ankle and wrist sensor sites in a pediatric A-T cohort or assessed longitudinal sensitivity of ankle-derived measures against clinician-rated scales in A-T.

*Added value of this study:* In an expanded pediatric cohort of 43 children with A-T and 50 controls followed for up to 5.8 years, we show that machine-learned composite scores derived from ankle and wrist accelerometer data collected during natural behavior at home correlate strongly with clinician-rated ataxia severity and caregiver-reported function, distinguish A-T from controls across age groups, and demonstrate high within-week reliability. The ankle composite captured disease progression with higher sensitivity than clinician-rated BARS, including in non-ambulatory individuals, whereas the wrist composite did not reach significance as a longitudinal measure. Both models identified a consistent motor signature of ataxia across upper and lower limbs.

*Implications of all the available evidence:* Passive at-home ankle accelerometry combined with submovement-based machine learning provides a sensitive, low-burden, and scalable measure of motor function and disease progression in A-T. This approach could complement clinician-rated scales as an objective outcome measure to increase clinical trial efficiency and provide early evidence for treatment efficacy. The low-cost single-sensor design supports geographically distributed trial enrollment, which is particularly relevant for rare diseases like A-T.

## Introduction

Sensitive, objective, and clinically meaningful outcome measures are needed to evaluate emerging therapies for neurodegenerative and neurodevelopmental conditions^1,2^. The development of disease-modifying therapies has accelerated for rare disorders with substantial unmet medical need^2^, but the tools currently used to assess motor function in trials and clinical care remain limited in their sensitivity to capture disease change and in their applicability across ages^3^. This challenge is amplified in conditions in which progressive neurodegeneration overlaps with ongoing motor development, where concurrent maturation can complicate the measurement of disease-related decline, particularly in early childhood^3^.

Ataxia-telangiectasia (A-T) is a rare autosomal recessive disorder caused by mutations in the ataxia-telangiectasia mutated *(ATM)* gene^4^, with progressive cerebellar atrophy giving rise to ataxia, dysarthria, oculomotor abnormalities, neuropathy, and extrapyramidal features. Motor signs typically emerge in early childhood and worsen through the first and second decades, with most individuals with classic A-T becoming wheelchair dependent by the early teenage years^5^, while milder phenotypes follow more variable trajectories^6^. Drug development in A-T is accelerating, with levacetylleucine^7^ recently receiving FDA approval for A-T, and other disease-modifying therapies under active clinical investigation, including triheptanoin^8^, nicotinamide riboside^9^, and individualized antisense oligonucleotide therapy targeting splice-altering ATM variants^10^. Despite this expanding landscape, sensitive pediatric-appropriate motor outcome measures remain a recognized bottleneck for A-T trial readiness^2^.

Clinician-performed ataxia rating scales, including the Brief Ataxia Rating Scale (BARS)^11^, the Scale for the Assessment and Rating of Ataxia (SARA)^12^, the International Cooperative Ataxia Rating Scale (ICARS)^13^, and the A-T Neurological Examination Scale Toolkit (A-T NEST)^14^, are the current standard for assessing motor severity in A-T. These scales are limited by rater subjectivity, low granularity of scoring, and insensitivity to small changes^15^. In pediatric A-T populations, an additional limitation is age dependence: ataxia rating scales yield non-zero baseline scores in healthy children up to approximately age eleven, constraining their ability to discriminate disease-related impairment from normal motor immaturity^3^. Caregiver-reported instruments such as the Caregiver Priorities and Child Health Index of Life with Disabilities (CPCHILD)^16^ provide complementary information about everyday functioning but share limitations of infrequent administration, subjectivity, and limited granularity^17^.

Wearable inertial sensors have emerged as a promising technology for the objective quantification of motor impairment in A-T and other pediatric and adult ataxias^18–20^. Task-based in-clinic protocols using accelerometers have distinguished ataxia from controls and correlated with rating-scale severity^21,22^, but share limitations with clinical scales in requiring participant cooperation, infrequent administration, and restricted applicability across the full age and severity range of pediatric populations. Task-free, at-home accelerometry, by contrast, captures continuous data during natural behavior without placing requirements on the participant^18^, supports inclusion of very young children and severely affected individuals, and has the potential to produce measures that closely reflect everyday motor function.

At-home wearable accelerometry has been explored in several ataxia populations including Friedreich’s ataxia^23^ and A-T^19,24^. We previously reported that wrist-derived submovement measures collected at home in children with A-T distinguish A-T from controls, correlate with BARS and CPCHILD, demonstrate high reliability, and show longitudinal change over a one-year interval^24^. Submovement-based approaches have also produced sensitive outcomes in adult cerebellar ataxias^20,21^, amyotrophic lateral sclerosis^25^, and prodromal Parkinson’s disease^26^, with composite machine-learned scores and ankle-based measures producing the most sensitive outcomes^20,21^; however, neither had yet been evaluated using at-home data in A-T.

To address these gaps, we test the hypothesis that submovement and activity-intensity features extracted from ankle and wrist accelerometer data collected continuously at home during natural behavior capture motor impairment and disease progression in A-T. In an expanded pediatric A-T cohort, we develop composite machine-learned severity scores from each sensor site, evaluate their relationships with clinician-rated motor severity (BARS) and caregiver-reported function (CPCHILD), characterize within-week reliability, examine age-stratified group separation, and compare the longitudinal sensitivity of ankle and wrist measures over a multi-year recording interval.

## Materials and methods

### Recruitment and consent

Participants with A-T and age- and sex-matched control participants took part in the study between January 2020 and December 2025. The study was approved by the Mass General Brigham Institutional Review Board (Protocol no. 2019P002752). Informed consent and, where applicable, assent were obtained from all participants and caregivers. Participants with A-T were recruited in collaboration with the Ataxia-Telangiectasia Children’s Project (A-TCP). Control participants included relatives of participants with A-T recruited through A-TCP. A detailed description of recruitment procedures is provided in the Supplementary Methods.

### Participants

Participants were classified by clinical A-T phenotype as classic A-T or mild A-T. Cross-sectional and longitudinal analyses were restricted to participants with classic A-T and age- and sex-matched controls. Four participants with mild A-T were retained in cross-sectional and longitudinal visualizations. The study began in January 2020 with wrist sensors only, and the ankle sensors were introduced in April 2022.

A total of 93 individuals were included: 43 children with classic A-T (22 female; median age 9.0 years; IQR 5.7-13.4; range 2.4-16.2; Table 1) and 50 controls (27 female; median age 8.8 years; IQR 5.6-12.3; range 2.1-17.5; Table 1). Sex was reported by caregivers at enrollment. The A-T cohort contributed 121 recording sessions, and controls contributed 128. Wrist data were available for all the sessions (A-T: 121; controls: 128), whereas ankle data were available for a subset (A-T: 83; controls: 87), owing to the later introduction of ankle sensors. Of the 38 A-T participants with ankle data (Table 1), 14 were non-ambulatory (wheelchair use >80% of the time), based on the caregiver-reported ambulation percentages. Control participants included the relatives of participants with A-T, a subset of whom had genetic confirmation that they carried one or no *ATM* mutations.

**Table 1.**
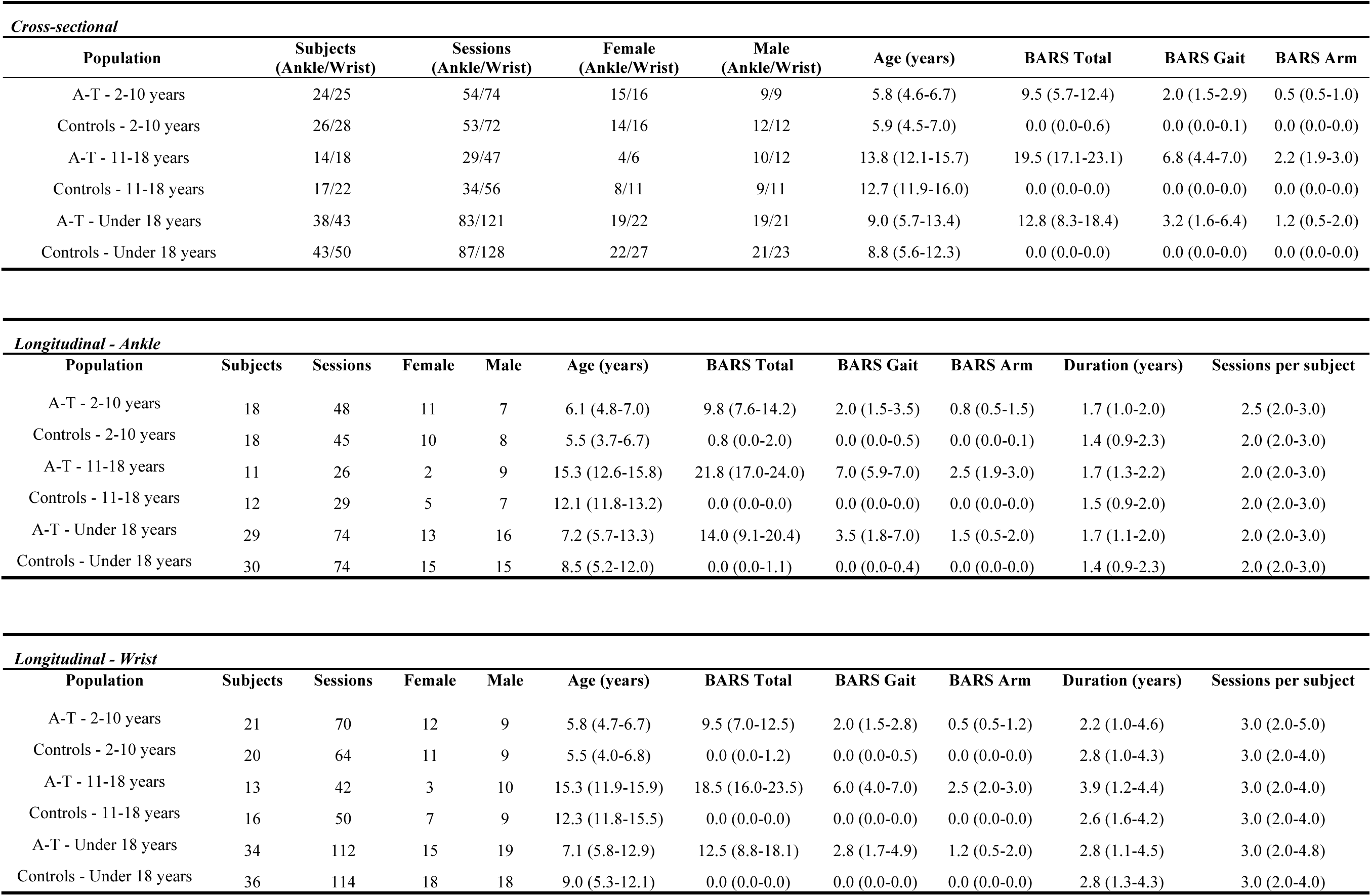
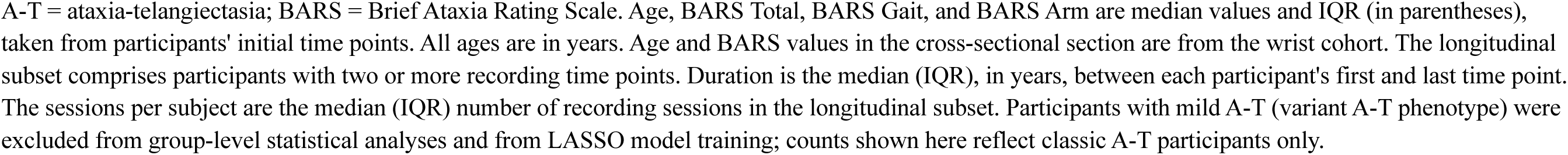
Summary of participant demographic and clinical information.

| ***Cross-sectional*** |  |  |  |  |  |  |  |  |  |  |
| **Population** | **Subjects (Ankle/Wrist)** | **Sessions (Ankle/Wrist)** | **Female (Ankle/Wrist)** | **Male (Ankle/Wrist)** | **Age (years)** | **BARS Total** | **BARS Gait** | **BARS Arm** |  |  |
|---|---|---|---|---|---|---|---|---|---|---|
| A-T - 2-10 years | 24/25 | 54/74 | 15/16 | 9/9 | 5.8 (4.6-6.7) | 9.5 (5.7-12.4) | 2.0 (1.5-2.9) | 0.5 (0.5-1.0) |  |  |
| Controls - 2-10 years | 26/28 | 53/72 | 14/16 | 12/12 | 5.9 (4.5-7.0) | 0.0 (0.0-0.6) | 0.0 (0.0-0.1) | 0.0 (0.0-0.0) |  |  |
| A-T - 11-18 years | 14/18 | 29/47 | 4/6 | 10/12 | 13.8 (12.1-15.7) | 19.5 (17.1-23.1) | 6.8 (4.4-7.0) | 2.2 (1.9-3.0) |  |  |
| Controls - 11-18 years | 17/22 | 34/56 | 8/11 | 9/11 | 12.7 (11.9-16.0) | 0.0 (0.0-0.0) | 0.0 (0.0-0.0) | 0.0 (0.0-0.0) |  |  |
| A-T - Under 18 years | 38/43 | 83/121 | 19/22 | 19/21 | 9.0 (5.7-13.4) | 12.8 (8.3-18.4) | 3.2 (1.6-6.4) | 1.2 (0.5-2.0) |  |  |
| Controls - Under 18 years | 43/50 | 87/128 | 22/27 | 21/23 | 8.8 (5.6-12.3) | 0.0 (0.0-0.0) | 0.0 (0.0-0.0) | 0.0 (0.0-0.0) |  |  |
| ***Longitudinal - Ankle*** |  |  |  |  |  |  |  |  |  |  |
| **Population** | **Subjects** | **Sessions** | **Female** | **Male** | **Age (years)** | **BARS Total** | **BARS Gait** | **BARS Arm** | **Duration (years)** | **Sessions per subject** |
| A-T - 2-10 years | 18 | 48 | 11 | 7 | 6.1 (4.8-7.0) | 9.8 (7.6-14.2) | 2.0 (1.5-3.5) | 0.8 (0.5-1.5) | 1.7 (1.0-2.0) | 2.5 (2.0-3.0) |
| Controls - 2-10 years | 18 | 45 | 10 | 8 | 5.5 (3.7-6.7) | 0.8 (0.0-2.0) | 0.0 (0.0-0.5) | 0.0 (0.0-0.1) | 1.4 (0.9-2.3) | 2.0 (2.0-3.0) |
| A-T - 11-18 years | 11 | 26 | 2 | 9 | 15.3 (12.6-15.8) | 21.8 (17.0-24.0) | 7.0 (5.9-7.0) | 2.5 (1.9-3.0) | 1.7 (1.3-2.2) | 2.0 (2.0-3.0) |
| Controls - 11-18 years | 12 | 29 | 5 | 7 | 12.1 (11.8-13.2) | 0.0 (0.0-0.0) | 0.0 (0.0-0.0) | 0.0 (0.0-0.0) | 1.5 (0.9-2.0) | 2.0 (2.0-3.0) |
| A-T - Under 18 years | 29 | 74 | 13 | 16 | 7.2 (5.7-13.3) | 14.0 (9.1-20.4) | 3.5 (1.8-7.0) | 1.5 (0.5-2.0) | 1.7 (1.1-2.0) | 2.0 (2.0-3.0) |
| Controls - Under 18 years | 30 | 74 | 15 | 15 | 8.5 (5.2-12.0) | 0.0 (0.0-1.1) | 0.0 (0.0-0.4) | 0.0 (0.0-0.0) | 1.4 (0.9-2.3) | 2.0 (2.0-3.0) |
| ***Longitudinal - Wrist*** |  |  |  |  |  |  |  |  |  |  |
| **Population** | **Subjects** | **Sessions** | **Female** | **Male** | **Age (years)** | **BARS Total** | **BARS Gait** | **BARS Arm** | **Duration (years)** | **Sessions per subject** |
| A-T - 2-10 years | 21 | 70 | 12 | 9 | 5.8 (4.7-6.7) | 9.5 (7.0-12.5) | 2.0 (1.5-2.8) | 0.5 (0.5-1.2) | 2.2 (1.0-4.6) | 3.0 (2.0-5.0) |
| Controls - 2-10 years | 20 | 64 | 11 | 9 | 5.5 (4.0-6.8) | 0.0 (0.0-1.2) | 0.0 (0.0-0.5) | 0.0 (0.0-0.0) | 2.8 (1.0-4.3) | 3.0 (2.0-4.0) |
| A-T - 11-18 years | 13 | 42 | 3 | 10 | 15.3 (11.9-15.9) | 18.5 (16.0-23.5) | 6.0 (4.0-7.0) | 2.5 (2.0-3.0) | 3.9 (1.2-4.4) | 3.0 (2.0-4.0) |
| Controls - 11-18 years | 16 | 50 | 7 | 9 | 12.3 (11.8-15.5) | 0.0 (0.0-0.0) | 0.0 (0.0-0.0) | 0.0 (0.0-0.0) | 2.6 (1.6-4.2) | 3.0 (2.0-4.0) |
| A-T - Under 18 years | 34 | 112 | 15 | 19 | 7.1 (5.8-12.9) | 12.5 (8.8-18.1) | 2.8 (1.7-4.9) | 1.2 (0.5-2.0) | 2.8 (1.1-4.5) | 3.0 (2.0-4.8) |
| Controls - Under 18 years | 36 | 114 | 18 | 18 | 9.0 (5.3-12.1) | 0.0 (0.0-0.0) | 0.0 (0.0-0.0) | 0.0 (0.0-0.0) | 2.8 (1.3-4.3) | 3.0 (2.0-4.0) |

A-T = ataxia-telangiectasia; BARS = Brief Ataxia Rating Scale. Age, BARS Total, BARS Gait, and BARS Arm are median values and IQR (in parentheses), taken from participants' initial time points. All ages are in years. Age and BARS values in the cross-sectional section are from the wrist cohort. The longitudinal subset comprises participants with two or more recording time points. Duration is the median (IQR), in years, between each participant's first and last time point. The sessions per subject are the median (IQR) number of recording sessions in the longitudinal subset. Participants with mild A-T (variant A-T phenotype) were excluded from group-level statistical analyses and from LASSO model training; counts shown here reflect classic A-T participants only.

Participants with two or more recording sessions at a sensor site were eligible for longitudinal analysis. The wrist longitudinal subset comprised 34 children with A-T and 36 controls (median three sessions per subject, IQR 2.0-4.8; range 2-6), and the ankle longitudinal subset comprised 29 children with A-T and 30 controls (median two sessions, IQR 2-3; range 2-4). Across the A-T longitudinal subset, the median follow-up duration was 2.8 years (IQR 1.1-4.5; range 0.8-5.8) for wrist and 1.7 years (IQR 1.1-2.0; range 0.8-3.5) for ankle.

### Virtual appointment and neurological assessment

At each time point, a study coordinator met with each participant and caregiver over video conference (in person in early 2020) to position and activate the GENEActiv wearable sensors and conduct a neurological assessment, which was video recorded for subsequent scoring. BARS was administered via video; a modified protocol (cBARS) was used for children under 13, developed for video-based scoring in children with A-T (details in Supplementary Methods). Scoring was performed by an ataxia-specialist neurologist. BARS scoring was available for 70 of 121 A-T sessions. Availability was affected by the transition from in-person to remote assessment during 2020-2021; analyses requiring BARS were restricted to sessions with available scores (see Supplementary Methods for year-by-year breakdown).

Caregiver-reported function was assessed using the CPCHILD questionnaire^16^, introduced into the study protocol in 2023. Analyses use the sum of Section 1 (Personal Care, nine items) and Section 2 (Positioning and Mobility, eight items), scored such that lower values indicate worse function. CPCHILD scores were available for 38 of 43 A-T participants and 40 of 50 controls.

### Wearable sensor data collection and processing

The GENEActiv actigraphy device measures triaxial acceleration at 100 Hz. Participants wore devices for seven days on the dominant wrist and, starting in April 2022, also on the dominant ankle. A minimum of five valid days of daytime data per sensor was required for inclusion (details in Supplementary Methods).

### Activity bouts and submovements

Activity intensity (AI), computed using the Activity Index^27^, summarized movement intensity based on the variance of acceleration across three axes^19^. Filtered accelerometer data were segmented into activity bouts at one-second resolution using an AI threshold to determine inactivity periods between bouts^24^. Using a previously validated approach^22,25^, we analyzed bouts of 4 to 18 seconds and decomposed the bouts into submovements (SMs). SMs were categorized by duration (short: 0.05-0.6 s; long: 0.6-5 s) and direction (primary/secondary)^24,25^. Each session yielded 85 features (details in Supplementary Methods).

### Composite severity models and single-feature analysis

We developed composite severity models by applying least absolute shrinkage and selection operator (LASSO) regression^28^ to features from each one-week recording session, generating a weighted linear combination to estimate severity. Separate models were trained for ankle (predicting BARS gait subscore) and wrist (predicting BARS total). Model training used 20-fold cross-validation with group-based splits to ensure no participant appeared in both training and test folds. Cross-sectional performance metrics were computed from out-of-fold predictions.

Single-feature analyses were performed on a pre-specified subset of seven features per sensor: the mean and standard deviation of distance, peak velocity, and peak acceleration for long-duration SMs in the secondary direction, plus AI entropy. The results are reported in Tables 2-6.

**Table 2.**
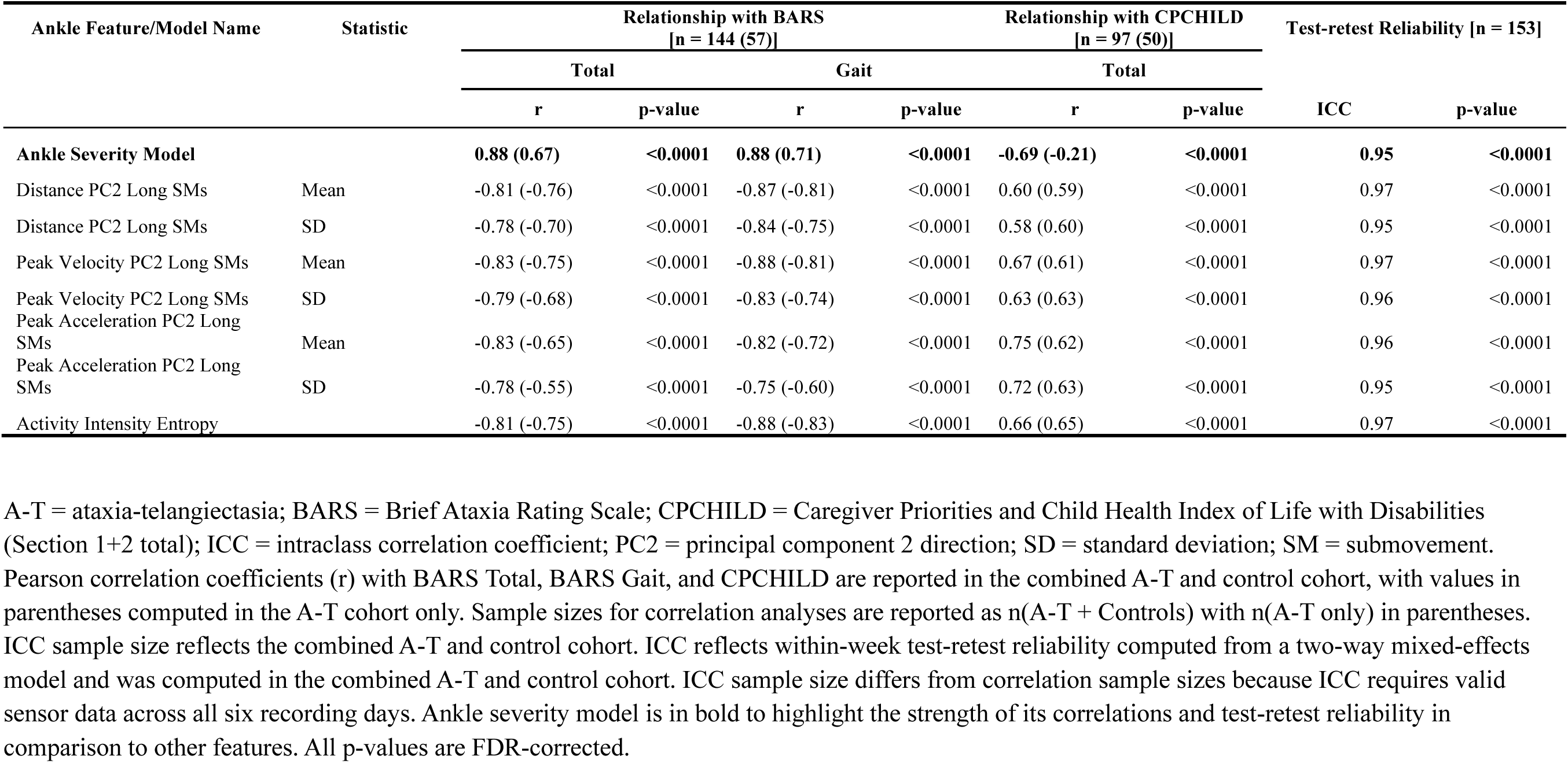
Cross-sectional relationship with clinical scales and within-week test-retest reliability for ankle sensor features and model.

### Statistical analyses

All statistical analyses were performed in Python (details in the Supplementary Methods). The Benjamini-Hochberg method was used to correct for multiple comparisons by controlling the false discovery rate (FDR); adjusted p < 0.05 was considered significant. Group differences between A-T and controls were assessed using Mann-Whitney U tests with Cohen’s d for effect size, stratified by age (2-10 and 11-18 years). Test-retest reliability was assessed using intraclass correlation coefficients (ICC; two-way mixed, consistency) computed from the first and second halves of each recording week.

Correlations between sensor-derived measures and clinical scores (BARS, CPCHILD) were assessed using Pearson correlation, computed for A-T and control participants combined (primary) and A-T participants alone (reported in parentheses in Tables 2 and 4).

Longitudinal sensitivity was assessed using the Mean-to-Standard Deviation Ratio (MSDR), defined as the ratio of the mean of per-subject progression slopes to the standard deviation of those slopes, with significance assessed using one-sample Wilcoxon signed-rank tests versus zero. To compare sensor and clinical scale sensitivity, per-subject slopes were computed for participants with longitudinal data for both sensor and clinical measures, and tested using Wilcoxon signed-rank tests versus zero, with Benjamini-Hochberg correction across all comparisons. Detailed statistical methodology is provided in the Supplementary Methods.

## Results

### Cross-sectional properties of ankle features

Predicted scores from the ankle severity model were strongly correlated with both the BARS gait subscore (r = 0.88; Fig. 1A and Table 2) and BARS total score (r = 0.88; Fig. 1B). The composite score was also correlated with caregiver-reported function on the CPCHILD total (r = -0.69). The model demonstrated excellent test-retest reliability (ICC = 0.95; Fig. 1C and Table 2) and distinguished A-T from control participants with a large effect size (Cohen’s d = 2.28; Fig. 1D and 1E and Table 3). Age-stratified analysis revealed a more pronounced group separation in the older cohort (ages 11-18; d = 3.48; Fig. 1E) compared to the younger cohort (ages 2-10; d = 2.01; Fig. 1E), consistent with the progressive nature of A-T (all Mann-Whitney U p < 0.0001).

**Figure 1.**
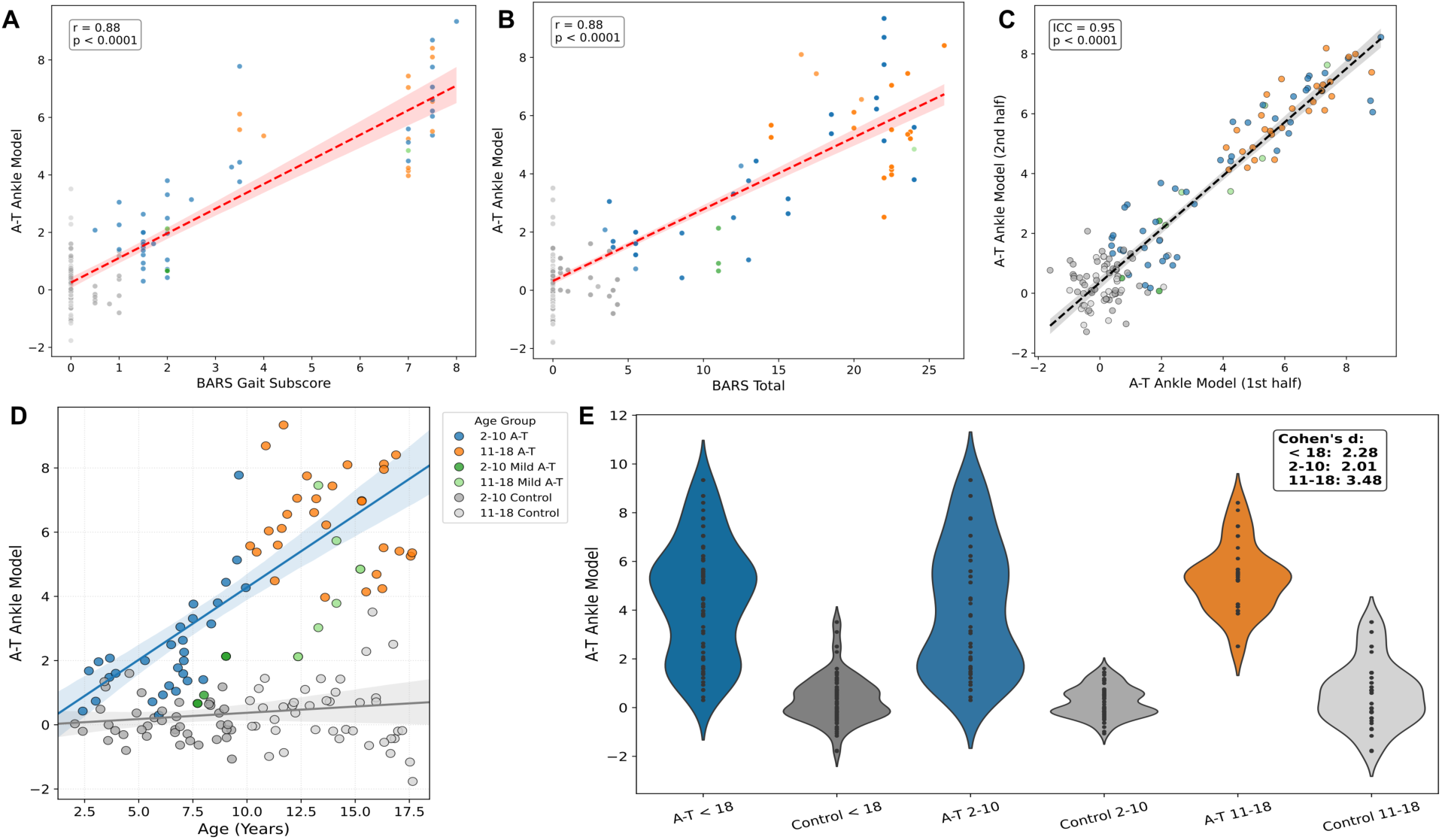
Cross-sectional properties of the ankle composite severity model and ankle features. **(A)** Scatter plot of out-of-fold predicted ankle composite severity scores (y-axis) versus BARS gait subscore (x-axis). Each point represents a single recording session. Points are colored by group (2-10 A-T, 11-18 A-T, 2-10 Mild A-T, 11-18 Mild A-T, 2-10 Control, 11-18 Control). A fitted linear regression line with 95% confidence interval is shown; Pearson r and p-value are annotated (r = 0.88, p < 0.0001). Sessions from participants with mild A-T (green) are shown for visualization only and were excluded from all statistical analyses. **(B)** Same as (A) but with BARS total score on the x-axis (r = 0.88, p < 0.0001). **(C)** Within-week test-retest reliability of the ankle composite score, comparing scores from days 1-3 (x-axis) versus days 4-6 (y-axis) of each recording session; ICC = 0.95, p < 0.0001. **(D)** Predicted ankle composite severity (y-axis) as a function of age (x-axis), with participants colored by group as in (A). Fitted linear regression lines with 95% confidence intervals are shown separately for the A-T and control groups. **(E)** Violin plots of predicted ankle composite scores stratified by group: A-T < 18, Control < 18, A-T 2-10, Control 2-10, A-T 11-18, Control 11-18. Individual session scores are overlaid as dots. Cohen’s d values are annotated (< 18: 2.28; 2-10: 2.01; 11-18: 3.48).

**Table 3.**
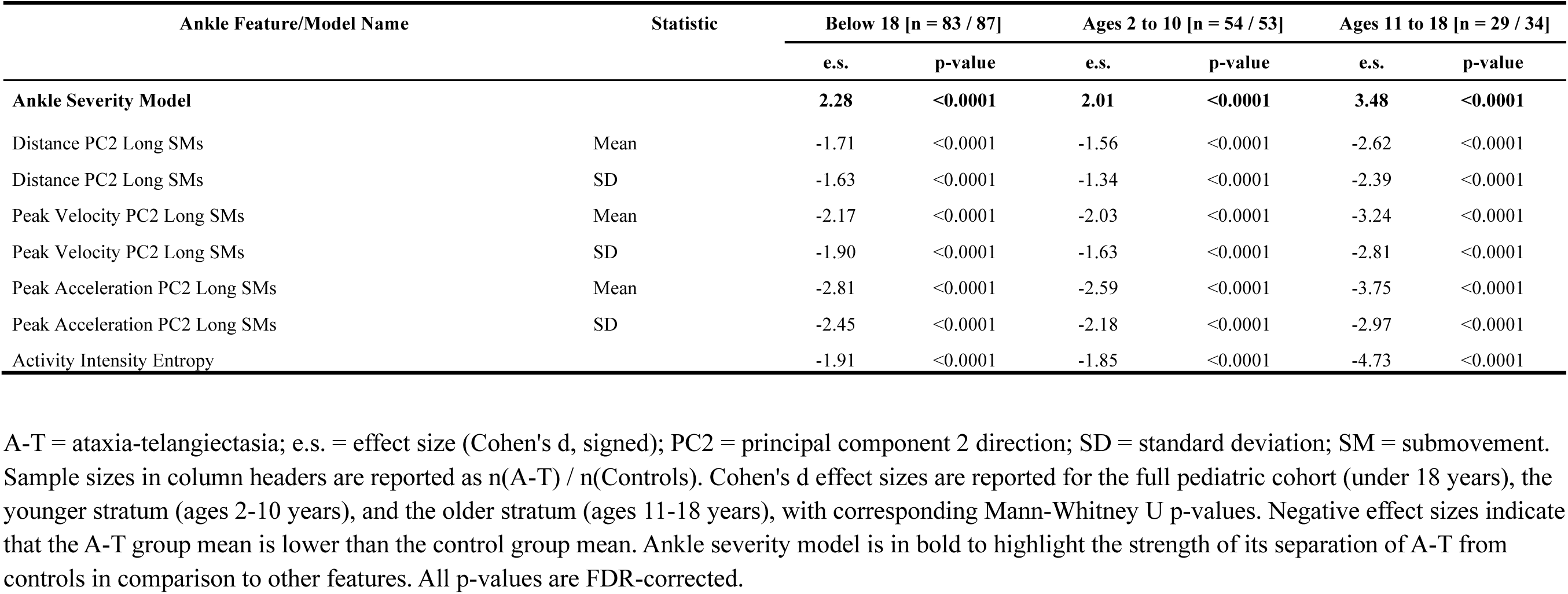
Discrimination between A-T and control participants for ankle sensor features and model.

Single-feature analysis on seven pre-specified ankle features confirmed these patterns. All features were significantly correlated with the BARS gait (|r| = 0.75-0.88; Table 2), BARS total (|r| = 0.78-0.83; all p < 0.0001, Table 2), and significantly associated with CPCHILD total (|r| = 0.58-0.75; Table 2). The features also demonstrated excellent test-retest reliability (ICC = 0.95-0.97; Table 2), and large group separation (|d| = 1.34-2.59 younger, 2.39-4.73 older; Table 3). All SM and AI features decreased with increasing disease severity, indicating smaller, slower, and less variable submovements.

### Cross-sectional properties of wrist features

The wrist severity model showed strong correlations with BARS total (r = 0.88; Fig. 2A), BARS arm subscore (r = 0.84; Fig. 2B), and CPCHILD (r = -0.78), with excellent reliability (ICC = 0.92; Fig. 2C) and large group separation (Cohen’s d = 2.98; Fig. 2D-E and Table 5). Age-stratified analysis showed strong group separation in the younger cohort (d = 2.68) and larger separation in the older cohort (d = 4.41; all p < 0.0001). Wrist composite scores increased with age in the A-T cohort while remaining stable in controls (Fig. 2D), paralleling the ankle findings.

**Figure 2.**
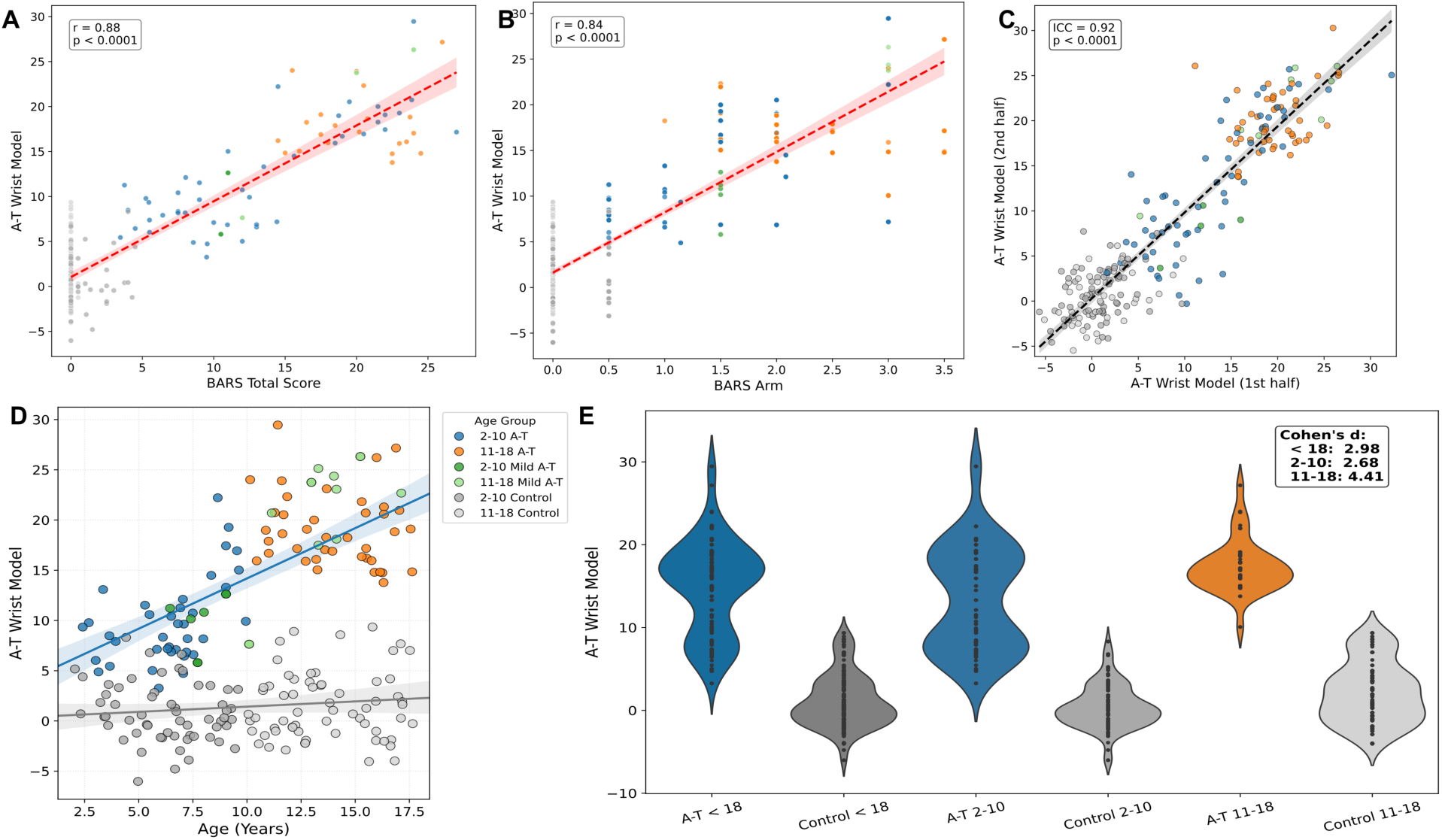
Cross-sectional properties of the wrist composite severity model and wrist features. **(A)** Scatter plot of out-of-fold predicted wrist composite severity scores (y-axis) versus BARS total score (x-axis). Points are colored by group (2-10 A-T, 11-18 A-T, 2-10 Mild A-T, 11-18 Mild A-T, 2-10 Control, 11-18 Control). A fitted linear regression line with 95% confidence interval is shown; Pearson r and p-value are annotated (r = 0.88, p < 0.0001). Sessions from participants with mild A-T (green) are shown for visualization only and were excluded from all statistical analyses. **(B)** Same as (A) but with BARS arm subscore on the x-axis (r = 0.84, p < 0.0001). **(C)** Within-week test-retest reliability of the wrist composite score, comparing scores from days 1-3 versus days 4-6 of each recording session; ICC = 0.92, p < 0.0001. **(D)** Predicted wrist composite severity as a function of age, with participants colored by group as in (A). Fitted linear regression lines with 95% confidence intervals are shown separately for the A-T and control groups. **(E)** Violin plots of predicted wrist composite scores stratified by group: A-T < 18, Control < 18, A-T 2-10, Control 2-10, A-T 11-18, Control 11-18. Cohen’s d values are annotated (< 18: 2.98; 2-10: 2.68; 11-18: 4.41).

Single-feature analysis on the corresponding seven wrist features showed significant correlations with BARS total (|r| = 0.60-0.83; Table 4) and BARS arm (|r| = 0.47-0.75), significant associations with CPCHILD (|r| = 0.54-0.73), high reliability (ICC = 0.90-0.95), and large effect sizes (|d| = 1.29-2.14 younger, 1.55-3.88 older; Table 5). As with the ankle, all SM and AI features decreased with increasing severity, indicating smaller, slower and less variable wrist movements (all p < 0.0001).

**Table 4.** Cross-sectional relationship with clinical scales and within-week test-retest reliability for wrist sensor features and model.

| Wrist Feature/Model Name | Statistic | Relationship with BARS<br>[n = 197 (70)] |  |  |  | Relationship with CPCHILD [n = 98 (50)] |  | Test-retest Reliability [n = 230] |  |
|---|---|---|---|---|---|---|---|---|---|
|  |  | Total |  | Arm |  | Total |  |  |  |
|  |  | r | p-value | r | p-value | r | p-value | ICC | p-value |
| **Wrist Severity Model** |  | **0.88 (0.53)** | **<0.0001** | **0.84 (0.46)** | **<0.0001** | **-0.78 (-0.36)** | **<0.0001** | **0.92** | **<0.0001** |
| Distance PC2 Long SMs | Mean | -0.83 (-0.71) | <0.0001 | -0.69 (-0.58) | <0.0001 | 0.70 (0.62) | <0.0001 | 0.94 | <0.0001 |
| Distance PC2 Long SMs | SD | -0.80 (-0.65) | <0.0001 | -0.66 (-0.53) | <0.0001 | 0.68 (0.61) | <0.0001 | 0.92 | <0.0001 |
| Peak Velocity PC2 Long SMs | Mean | -0.82 (-0.68) | <0.0001 | -0.68 (-0.56) | <0.0001 | 0.73 (0.62) | <0.0001 | 0.94 | <0.0001 |
| Peak Velocity PC2 Long SMs | SD | -0.78 (-0.62) | <0.0001 | -0.64 (-0.51) | <0.0001 | 0.68 (0.60) | <0.0001 | 0.93 | <0.0001 |
| Peak Acceleration PC2 Long SMs | Mean | -0.71 (-0.45) | <0.0001 | -0.55 (-0.38) | <0.0001 | 0.71 (0.54) | <0.0001 | 0.95 | <0.0001 |
| Peak Acceleration PC2 Long SMs | SD | -0.60 (-0.34) | <0.0001 | -0.47 (-0.31) | <0.0001 | 0.54 (0.52) | <0.0001 | 0.90 | <0.0001 |
| Activity Intensity Entropy |  | -0.82 (-0.71) | <0.0001 | -0.75 (-0.68) | <0.0001 | 0.64 (0.68) | <0.0001 | 0.95 | <0.0001 |

A-T = ataxia-telangiectasia; BARS = Brief Ataxia Rating Scale; CPCHILD = Caregiver Priorities and Child Health Index of Life with Disabilities (Section 1+2 total); ICC = intraclass correlation coefficient; PC2 = principal component 2 direction; SD = standard deviation; SM = submovement. Pearson correlation coefficients (r) with BARS Total, BARS Arm, and CPCHILD are reported in the combined A-T and control cohort, with values in parentheses computed in the A-T cohort only. Sample sizes for correlation analyses are reported as n(A-T + Controls) with n(A-T only) in parentheses. ICC sample size reflects the combined A-T and control cohort. ICC reflects within-week test-retest reliability computed from a two-way mixed-effects model and was computed in the combined A-T and control cohort. ICC sample size differs from correlation sample sizes because ICC requires valid sensor data across all six recording days. Wrist severity model is in bold to highlight the strength of its correlations and test-retest reliability in comparison to other features. All p-values are FDR-corrected.

**Table 5.** Discrimination between A-T and control participants for wrist sensor features and model.

| **Wrist Feature/Model Name** | **Statistic** | **Below 18 [n = 121 / 128]** |  | **Ages 2 to 10 [n = 74 / 72]** |  | **Ages 11 to 18 [n = 47 / 56]** |  |
|---|---|---|---|---|---|---|---|
|  |  | **e.s.** | **p-value** | **e.s.** | **p-value** | **e.s.** | **p-value** |
| **Wrist Severity Model** |  | **2.98** | **<0.0001** | **2.68** | **<0.0001** | **4.41** | **<0.0001** |
| Distance PC2 Long SMs | Mean | -2.06 | <0.0001 | -1.94 | <0.0001 | -3.87 | <0.0001 |
| Distance PC2 Long SMs | SD | -2.11 | <0.0001 | -1.98 | <0.0001 | -3.82 | <0.0001 |
| Peak Velocity PC2 Long SMs | Mean | -2.25 | <0.0001 | -2.14 | <0.0001 | -3.88 | <0.0001 |
| Peak Velocity PC2 Long SMs | SD | -2.14 | <0.0001 | -2.00 | <0.0001 | -3.45 | <0.0001 |
| Peak Acceleration PC2 Long SMs | Mean | -2.24 | <0.0001 | -2.10 | <0.0001 | -3.04 | <0.0001 |
| Peak Acceleration PC2 Long SMs | SD | -1.47 | <0.0001 | -1.29 | <0.0001 | -1.55 | <0.0001 |
| Activity Intensity Entropy |  | -1.66 | <0.0001 | -1.59 | <0.0001 | -2.99 | <0.0001 |

A-T = ataxia-telangiectasia; e.s. = effect size (Cohen's d, signed); PC2 = principal component 2 direction; SD = standard deviation; SM = submovement. Sample sizes in column headers are reported as n(A-T) / n(Controls). Cohen's d effect sizes are reported for the full pediatric cohort (under 18 years), the younger stratum (ages 2-10 years), and the older stratum (ages 11-18 years), with corresponding Mann-Whitney U p-values. Negative effect sizes indicate that the A-T group mean is lower than the control group mean. Wrist severity model is in bold to highlight the strength of its separation of A-T from controls in comparison to other features. All p-values are FDR-corrected.

### Longitudinal analysis

The ankle severity model demonstrated strong sensitivity to disease progression in A-T participants with longitudinal recordings (n = 29; MSDR = 0.98, p < 0.0001; Table 6 and Fig. 3A). Including participants with mild A-T, individuals in both ambulatory (15/18 positive slopes) and non-ambulatory (13/14) categories showed progression, supporting the ability to detect change across stages of disease. All seven individual ankle features showed significant change over time (MSDR = -0.41 to -0.92; Table 6), with mean SM distance (MSDR = -0.92) and mean peak velocity (MSDR = -0.91) demonstrating the highest sensitivity, indicating that ankle SMs became progressively smaller and slower. Controls exhibited stable predicted severity (Fig. 3A).

**Table 6.** Sensitivity to disease progression for ankle and wrist sensor features and models.

| Feature/Model Name | Statistic | Ankle Sensor (n = 29) |  | Wrist Sensor (n = 34) |  |
|---|---|---|---|---|---|
|  |  | e.s. | p-value | e.s. | p-value |
| **Ankle Severity Model** |  | **0.98** | **<0.0001** | - | - |
| Wrist Severity Model |  | - | - | 0.21 | 0.45 |
| Distance PC2 Long SMs | Mean | -0.92 | <0.0001 | -0.58 | 0.00063 |
| Distance PC2 Long SMs | SD | -0.62 | 0.0041 | -0.50 | 0.0044 |
| Peak Velocity PC2 Long SMs | Mean | -0.91 | <0.0001 | -0.53 | 0.0023 |
| Peak Velocity PC2 Long SMs | SD | -0.63 | 0.0038 | -0.37 | 0.028 |
| Peak Acceleration PC2 Long SMs | Mean | -0.65 | 0.00024 | -0.35 | 0.060 |
| Peak Acceleration PC2 Long SMs | SD | -0.41 | 0.023 | -0.09 | 0.61 |
| Activity Intensity Entropy |  | -0.74 | 0.00045 | -0.30 | 0.010 |

A-T = ataxia-telangiectasia; e.s. = effect size (MSDR); MSDR = mean-to-standard-deviation ratio; PC2 = principal component 2 direction; SD = standard deviation; SM = submovement. MSDR values are reported with p-values from one-sample Wilcoxon signed-rank tests against zero. Negative effect sizes indicate a decrease in the feature value over time. Ankle analyses include 29 participants with A-T; wrist analyses include 34 participants with A-T (full longitudinal subset; no BARS matching). Composite models are reported only for the sensor on which the model was trained (- = not applicable). Ankle severity model is in bold to highlight its strong sensitivity to disease progression in comparison to other features. All p-values are FDR-corrected.

**Figure 3.**
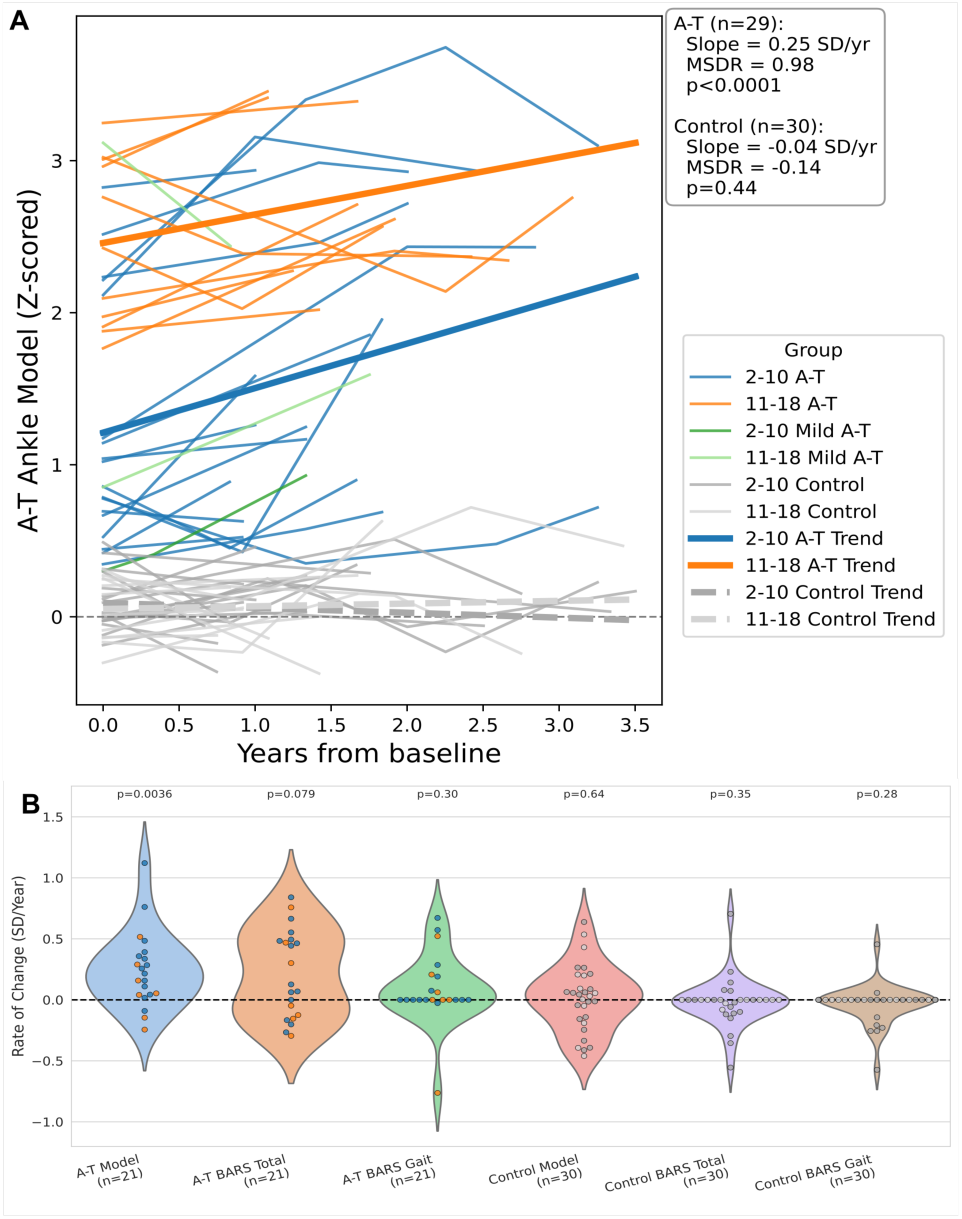
Longitudinal change in ankle composite scores. **(A)** Per-subject longitudinal trajectories of predicted ankle composite scores (z-scored) over years from baseline, in A-T participants (n = 29; stratified by age 2-10 and 11-18 years) and control participants (n = 30; stratified by age 2-10 and 11-18 years). Individual subject trajectories are shown as thin lines; bold trend lines depict the group-level trajectory for A-T (solid) and controls (dashed). Trajectories from participants with mild A-T are shown for visualization only and were excluded from statistical analyses. Per-subject slope statistics are annotated: A-T mean slope = 0.25 SD/year (MSDR = 0.98, p < 0.0001); control mean slope = −0.04 SD/year (MSDR = −0.14, p = 0.44). MSDR denotes the mean-to-standard-deviation ratio of per-subject slopes. **(B)** Per-subject rates of change (slopes) for the ankle composite model, BARS Total, and BARS Gait in the subset of A-T participants (n = 21) and controls (n = 30) with longitudinal data for all three measures. Points are colored by age subgroup (2-10 and 11-18 years) for A-T and controls. P-values from one-sample Wilcoxon signed-rank tests against zero are shown above each violin, after Benjamini-Hochberg FDR correction within the panel.

The wrist composite model (n = 34) did not demonstrate significant longitudinal sensitivity. Among individual wrist features, five of seven reached significance (MSDR = -0.30 to -0.58; Table 6), with mean SM distance (MSDR = -0.58) and mean peak velocity (MSDR = -0.53) showing the strongest signals. Thus, wrist SMs also became smaller and slower over time, but with lower MSDRs than the ankle.

To compare the longitudinal sensitivity of ankle-derived measures with clinician-rated scales, we computed per-subject rate-of-change distributions for participants with longitudinal data for both sensor and clinical measures (n = 21 A-T, n = 30 controls; Fig. 3B). This subset is smaller than the full ankle longitudinal cohort (Fig. 3A; n = 29 A-T, n = 30 controls) because some participants with longitudinal sensor data did not have BARS scored at two or more time points. Ankle composite slopes were significantly greater than zero in A-T (Wilcoxon signed-rank, p = 0.0036), whereas BARS total (p = 0.079) and BARS gait (p = 0.30) did not reach significance. Control slope distributions did not differ from zero for any measure. P-values were jointly corrected (Benjamini-Hochberg) across all measures and groups.

### Model feature analysis

To understand which individual features were most salient, we identified the five features with the largest absolute coefficients in each model. The ankle model was led by AI entropy (largest coefficient), the mean of the first shape component of long-duration SMs in the secondary direction, variability in short-duration SM peak velocity (primary direction) and peak acceleration (secondary direction), and the median of the AI histogram (see reference ^26^ and the Supplementary Methods for a detailed description of the features). The wrist model shared AI entropy and the first shape component as its top two features, with normalized jerk of long-duration SMs in primary direction, and direction explained at high and moderate activity intensity rounding out the top five. The overlap in leading features suggests both models capture a consistent motor signature of ataxia across upper and lower limbs.

## Discussion

We have shown that data from relatively low-cost sensors worn on the ankle and wrist at home during natural behavior produce reliable and sensitive measures of motor function that are interpretable and align with clinician assessment and caregiver report in ataxia-telangiectasia. SM and activity features extracted from the ankle and wrist distinguished individuals with A-T from controls, correlated with clinician-rated ataxia severity and caregiver-reported function, and demonstrated high within-week reliability. Notably, the ankle-derived composite measure captured disease progression sensitively over time, whereas the wrist-derived composite measure did not, although the majority of individual wrist features showed significant progression with smaller effect sizes. Together, these findings support the use of passive wearable sensing, particularly at the ankle location, as an ecologically valid and scalable approach for sensitively monitoring motor function in A-T.

Both the ankle and the wrist composite scores showed similarly strong associations with overall ataxia severity (BARS total) and caregiver-reported function on CPCHILD, indicating that the sensor-derived measures from both sites capture motor impairment that aligns with clinician and caregiver assessments. At the individual feature level, ankle features were more strongly associated with the BARS gait subscore than wrist features were with the BARS arm subscore, which may reflect the greater granularity of the BARS gait subscore relative to the arm subscore, or the fact that upper-limb movement encompasses a broad range of activities not fully captured by the BARS arm subscore.

Both ankle- and wrist-based composite scores increased with age in the A-T cohort while remaining stable in controls (Fig. 1D, 2D), indicating that the models capture disease-related motor decline rather than typical developmental change. This is an important distinction because overlapping motor development and neurodegeneration in early childhood can mask decline on clinician-rated scales^3^. The models significantly separated A-T from controls even in the younger cohort, supporting their potential for detecting motor impairment at ages when clinical scales are less sensitive.

There was substantial overlap in the most salient features between ankle and wrist models, including AI entropy, where lower values reflect reduced range of movement intensities, and the first shape component of long-duration SMs in the secondary direction, where reduced values indicate less smooth and less bell-shaped velocity profiles. This suggests a consistent motor signature of ataxia across upper and lower limbs. The consistent negative relationship between SM features and BARS indicates movements become smaller, slower, and less variable with increasing severity, consistent with progressive movement decomposition reported in prior wearable^20,24^ and task-based ataxia^29^ studies.

The ankle composite score detected disease progression with higher sensitivity than clinician-rated BARS in the matched longitudinal subset, while controls remained stable over the same period. This advantage likely reflects the continuous, objective nature of sensor measurement, which aggregates information from many hours of natural behavior compared with brief clinical assessments at discrete time points. The strongest longitudinal signals arose from SM distance and peak velocity, consistent with progressive movement decomposition in ataxia, where movements become smaller and slower over time. Notably, progression was evident even among individuals who were no longer able to ambulate independently, indicating that the ankle signal captures disease-related motor change beyond gait deterioration, consistent with lower-limb impairment being a prominent and progressive feature of A-T.

The wrist composite model did not reach significance as a longitudinal measure, although the majority of individual wrist features demonstrated significant progression with smaller effect sizes. Because the composite was trained to predict cross-sectional severity, its feature weights may not optimally capture within-subject change over time; a model trained with a longitudinal learning objective may be needed to translate feature-level wrist sensitivity into a composite progression measure^25^. The ankle composite captured significant progression under a cross-sectional training objective, and a change-optimized approach could further strengthen its sensitivity.

The strong cross-sectional and longitudinal sensitivity of the ankle composite model is consistent with recent ankle-based wearable work in adult cerebellar ataxias^20,30^, which reported strong correlations with clinical scores and longitudinal progression in spinocerebellar ataxias and multiple system atrophy, cerebellar type (MSA-C). The asymmetry between the ankle and wrist longitudinal sensitivity that we report in A-T also resembles findings in that adult ataxia population, where ankle composite progression signals were among the most pronounced. More broadly, submovement-based approaches have demonstrated broad applicability across pediatric^24,29^ and adult ataxia populations^20–22^ as well as other neurological populations including amyotrophic lateral sclerosis (ALS)^25^ and prodromal Parkinson’s disease^26^.

The ecological validity, reliability, and longitudinal sensitivity of the ankle composite measure suggest potential clinical applications. As an outcome measure in A-T clinical trials, an ankle-based composite could provide a continuous, objective signal that complements clinician-rated scales, particularly in age groups where rating scales are limited by motor immaturity or floor effects. The low-cost single-sensor design reduces examiner burden and supports geographically distributed enrollment, both relevant for a rare disease. The measure does not require a structured walking task and captures progression in non-ambulatory individuals.

There are some limitations. Although the cohort is large for a rare disease, the absolute sample size remains modest, particularly for the longitudinal subset (ankle n = 29; wrist n = 34), limiting power for subgroup analyses. BARS and CPCHILD scores were not available for all sessions, in part due to the transition from in-person to remote assessment; however, the available scores spanned the severity spectrum and provided sufficient data for evaluating associations. Broader availability of clinical scores would expand training data and could further improve model performance. The clinical assessments used for comparison share the limitations of clinician-rated and caregiver-rated instruments, including subjectivity, low granularity, and ceiling/floor effects.

In summary, passive at-home accelerometry from the ankle and wrist produces reliable and sensitive measures of motor function that are interpretable and aligned with clinical assessments in ataxia-telangiectasia. This study supports the use of low-cost, low-burden wearable sensors as outcome measures in A-T clinical care and clinical trials, offering the potential to provide more sensitive and more accessible motor assessments compared with conventional rating scales.

## Data Availability

De-identified participant data and a data dictionary will be made available to qualified investigators upon reasonable request to the corresponding author.

## Contributors

DK and ASG contributed to the conception and design of the study. ART, AL, SR, and JT contributed to participant recruitment. ART and AL contributed to data acquisition, coordinated study visits, and administered the BARS assessments. FH scored the video-recorded BARS assessments. DK performed the data analysis. ASG contributed to funding acquisition, supervision, and validation of the results. ASG and FH critically reviewed the manuscript. DK and ASG drafted the manuscript and prepared the figures. DK and ASG accessed and verified the underlying data. All authors had full access to all the data in the study, reviewed and approved the final manuscript, and had final responsibility for the decision to submit for publication.

## Acknowledgments

We thank the participants and their families for their time and dedication to this research, and the A-T Children’s Project for its partnership in participant recruitment. This work was supported by the U.S. National Institutes of Health (R01 NS134597), the A-T Children’s Project, and Insmed.

## AI Declaration

The authors used AI tools to suggest edits to condense the manuscript and improve clarity, identify grammatical errors, and identify additional literature of relevance.

## References

1 Gupta AS. Digital phenotyping in clinical neurology. Semin Neurol 2022; 42: 48–59.

2 Kuhn K, Lederman HM, McGrath-Morrow SA. Ataxia-telangiectasia clinical trial landscape and the obstacles to overcome. Expert Opin Investig Drugs 2023; 32: 693–704.

3 Brandsma R, Spits AH, Kuiper MJ, et al. Ataxia rating scales are age-dependent in healthy children. Dev Med Child Neurol 2014; 56: 556–63.

4 Rothblum-Oviatt C, Wright J, Lefton-Greif MA, McGrath-Morrow SA, Crawford TO, Lederman HM. Ataxia telangiectasia: a review. Orphanet J Rare Dis 2016; 11: 159.

5 Petley E, Yule A, Alexander S, Ojha S, Whitehouse WP. The natural history of ataxia-telangiectasia (A-T): A systematic review. PLoS One 2022; 17: e0264177.

6 van Os NJH, Hensiek A, van Gaalen J, et al. Trajectories of motor abnormalities in milder phenotypes of ataxia telangiectasia. Neurology 2019; 92: e19–29.

7 Martakis K, Bremova-Ertl T, Bolton C, et al. Safety and efficacy of levacetylleucine in ataxia-telangiectasia: a phase 3, randomised, double-blind, placebo-controlled crossover trial. Lancet Neurol 2026; 25: 633–44.

8 Onodera O. Triheptanoin treatment in ataxia-telangiectasia: the significance of innovative clinical trials targeting mitochondrial dysfunction. EBioMedicine 2025; 119: 105882.

9 Presterud R, Deng WH, Wennerström AB, et al. Long-term nicotinamide riboside use improves coordination and eye movements in ataxia telangiectasia. Mov Disord 2024; 39: 360–9.

10 Kim J, Woo S, de Gusmao CM, et al. A framework for individualized splice-switching oligonucleotide therapy. Nature 2023; 619: 828–36.

11 Schmahmann JD, Gardner R, MacMore J, Vangel MG. Development of a brief ataxia rating scale (BARS) based on a modified form of the ICARS. Mov Disord 2009; 24: 1820–8.

12 Schmitz-Hübsch T, du Montcel ST, Baliko L, et al. Scale for the assessment and rating of ataxia: development of a new clinical scale. Neurology 2006; 66: 1717–20.

13 Trouillas P, Takayanagi T, Hallett M, et al. International Cooperative Ataxia Rating Scale for pharmacological assessment of the cerebellar syndrome. The Ataxia Neuropharmacology Committee of the World Federation of Neurology. J Neurol Sci 1997; 145: 205–11.

14 Crawford TO, Mandir AS, Lefton-Greif MA, et al. Quantitative neurologic assessment of ataxia-telangiectasia. Neurology 2000; 54: 1505–9.

15 Klockgether T, Synofzik M, AGI working group on COAs and Registries. Consensus recommendations for clinical outcome assessments and registry development in ataxias: Ataxia Global Initiative (AGI) working group expert guidance. Cerebellum 2024; 23: 924–30.

16 Narayanan UG, Fehlings D, Weir S, Knights S, Kiran S, Campbell K. Initial development and validation of the Caregiver Priorities and Child Health Index of Life with Disabilities (CPCHILD). Dev Med Child Neurol 2006; 48: 804–12.

17 Cella D, Hahn E, Jensen S, et al. Patient-reported outcomes in performance measurement. RTI Press, 2015.

18 Ilg W, Seemann J, Giese M, et al. Real-life gait assessment in degenerative cerebellar ataxia: Toward ecologically valid biomarkers. Neurology 2020; 95: e1199–210.

19 Khan NC, Pandey V, Gajos KZ, Gupta AS. Free-living motor activity monitoring in ataxia-telangiectasia. Cerebellum 2022; 21: 368–79.

20 Manohar R, Yang FX, Stephen CD, Schmahmann JD, Eklund NM, Gupta AS. At-home wearables and machine learning capture motor impairment and progression in adult ataxias. Brain 2025; 148: 3623–34.

21 Patel S, Oubre B, Stephen CD, Schmahmann JD, Gupta AS. Submovements derived from wearable sensors capture ataxia severity and differ across motor tasks and directions of motion. Cerebellum 2025; 24: 156.

22 Oubre B, Daneault J-F, Whritenour K, et al. Decomposition of reaching movements enables detection and measurement of ataxia. Cerebellum 2021; 20: 811–22.

23 Mishra RK, Nunes AS, Enriquez A, et al. At-home wearable-based monitoring predicts clinical measures and biological biomarkers of disease severity in Friedreich’s Ataxia. Commun Med (Lond) 2024; 4: 217.

24 Gupta AS, Luddy AC, Khan NC, Reiling S, Thornton JK. Real-life wrist movement patterns capture motor impairment in individuals with ataxia-telangiectasia. Cerebellum 2023; 22: 261–71.

25 Gupta AS, Patel S, Premasiri A, Vieira F. At-home wearables and machine learning sensitively capture disease progression in amyotrophic lateral sclerosis. Nat Commun 2023; 14: 5080.

26 Gupta AS, Patel S. Wrist accelerometry and machine learning sensitively capture disease progression in prodromal Parkinson’s disease. NPJ Parkinsons Dis 2025; 11: 171.

27 Bai J, Di C, Xiao L, et al. An activity index for raw accelerometry data and its comparison with other activity metrics. PLoS One 2016; 11: e0160644.

28 Tibshirani R. Regression shrinkage and selection via the lasso. J R Stat Soc Series B Stat Methodol 1996; 58: 267–88.

29 Lee J, Oubre B, Daneault J-F, Lee SI, Gupta AS. Estimation of ataxia severity in children with ataxia-telangiectasia using ankle-worn sensors. J Neurol 2023; 270: 5097–101.

30 Seemann J, Daghsen L, Cazier M, et al. Digital gait measures capture 1-year progression in early-stage spinocerebellar ataxia type 2. Mov Disord 2024; 39: 788–97.

