## Supplementary Methods for "At-home ankle and wrist accelerometry and machine learning capture motor impairment and disease progression in ataxia-telangiectasia"

### Recruitment and consent

Study participants with A-T were recruited in collaboration with the Ataxia-Telangiectasia Children's Project (A-TCP). From the A-TCP database, the study team contacted parents and participants by email with a description of the study procedures and scheduled a call to complete consent. Study participants met the following criteria: (i) a clinical and/or molecular diagnosis of A-T; (ii) age 18 months or older; (iii) ability to wear ankle and wrist accelerometers continuously for one week; and (iv) ability to participate in remote study assessments by video conference. Participants with another neurological condition that could affect their movement or an unstable medical illness were excluded. Age- and sex-matched relatives of participants with A-T were recruited as control participants through the same A-TCP partnership and met the same age, wear, technology, and consent criteria; the absence of any other neurological or medical condition was the principal screening criterion for controls. The study protocol initially scheduled repeat visits at 12-month intervals; the protocol was subsequently revised to 6-month intervals to better characterize motor change over time, and some families retained 12-month intervals owing to scheduling constraints.

### Virtual appointment and neurological assessment

BARS administration followed a modified protocol for children under age 13 (cBARS) developed for video-based scoring in children with A-T. The examination order followed a standardized sequence: speech, finger-to-nose, oculomotor, stance, gait, and heel-to-shin, performed under the video coordination of the study coordinator and (for younger participants) the caregiver. Age-appropriate supplies were shipped to each participant in advance to support the in-home examination: for participants aged 1-3 years, the package included two blocks, two animal faces, and a drawing sheet with instructions; for participants aged 4-6 years, the package additionally included a marker; and for participants aged 7-12 years, the package included the drawing sheet (and animal faces for ages 7-9). The drawing component of the examination did not need to be completed during the video appointment and was returned by photograph, scan, or hard-copy mail. For participants aged 12 years and younger, the cBARS scoring instrument was completed in addition to the standard BARS scoring instrument; for participants aged 13 years and older, only the standard BARS half-point scoring was completed.

Scoring was performed by an ataxia-specialist neurologist from the recordings of each video-recorded assessment. The rater was blinded to the participant group. Where individual BARS subscores could not be assigned for reasons including poor task performance, the rater left a comment indicating the reason. The clinical assessments were initially conducted in person in early 2020 during the annual A-T fundraiser in Orlando, Florida, and all 11 of the 2020 sessions had associated clinical scores. Due to the COVID-19 pandemic, in-person evaluations were not possible in 2021 and only 1 out of 24 sessions had clinical scores. In 2022, we began to conduct virtual assessments over Zoom. This generally worked well; however, there were occasional logistical, internet connectivity, and pediatric participation challenges. From 2022 onward, 58 out of 86 sessions from A-T participants had associated clinical scores.

Wearable sensor data collection and processing

This study used the GENEActiv actigraphy device[1] (ActivInsights Ltd, Cambridge, UK) that measures triaxial acceleration at 100 Hz with a MEMS sensor (range: ±8 g; resolution: 12-bit). The minimum wear time threshold was set to 5 hours per day and a minimum of 5 days. Data analysis commenced from the first full 24-hour recording following device placement, resulting in six consecutive 24-hour periods of data for most weeks of wear. We term this a "session". Each participant's raw accelerometer data were manually partitioned into day and night segments based on their activity levels as previously described[2]. In this work, the analyses focused on the daytime data. Gravity and high-frequency noise were removed from the daytime data using a sixth-order Butterworth filter with cutoff frequencies of 0.1 and 20 Hz.

Feature enumeration

Each session yielded 85 features: 40 kinematic features from SMs (5 properties - distance, duration, peak velocity, peak acceleration, and jerk, each summarized by mean and standard deviation for each direction of motion and each duration class); 30 shape features from long-duration SMs (3 statistics applied to each of 5 principal components of the velocity-time profile, for each direction of motion); 10 AI features[2] (mean, median, and entropy of the daytime AI distribution; mean bout AI; percentage of daytime at low, moderate and high AI; and the percentage of acceleration variance explained by the principal movement direction within low-, moderate- and high-AI 1-s time bins); 4 activity-bout features (mean and standard deviation of jerk and acceleration); and 1 spectral feature (total power in the 0.1-5 Hz frequency range)[2].

Detailed statistical methodology

The Benjamini-Hochberg method was used to control the false discovery rate (FDR)[3] and to adjust the p-values for multiple comparisons in all the statistical tests performed, and the corrected p-values < 0.05 were considered significant. We employed the Mann-Whitney U test[4] to determine feature differences between the A-T and control groups, with Cohen's d to measure the effect size. The effect sizes are signed, where negative values indicate the mean of the A-T group was smaller than that of the control group. The reliability of the ankle and wrist sensor measurements was assessed by computing the Intraclass Correlation Coefficient (ICC)[5]. Each session was divided into two sub-sessions, days 1 to 3 and days 4 to 6, and the reliability was computed using a two-way mixed effects model (ICC3)[6] for consistency between the features of the sub-sessions. ICC was computed in the combined A-T and control cohort and was restricted to subjects with valid data across all six recording days; the resulting sample size, therefore, differs from other cross-sectional analyses and is reported in the corresponding table. To validate the clinical relevance of the ankle and wrist digital features, their relationship with the Brief Ataxia Rating Scale (BARS) was examined using both A-T and control participants. Pearson's correlation was used to measure the association between the movement metrics and the clinical severity. Pearson correlations were computed in the combined A-T and control cohort (primary value) and additionally restricted to the A-T cohort only (reported in parentheses). P-values reported in correlation tables reflect the combined-cohort correlation after Benjamini-Hochberg FDR correction. Feature-specific correlations were performed based on the anatomical relevance; wrist-worn sensors were correlated with upper-limb clinical scores, while the ankle-worn sensors were correlated with gait subscores, in addition to the correlation analysis of both ankle and wrist sensors with the BARS total score. To further validate the clinical relevance of the composite models, predicted severity scores were correlated with the caregiver-reported CPCHILD total, as defined in the main text. The ability of movement features to detect disease progression over time was evaluated through per-subject progression rates, computed as the linear regression slope of each feature on time (in years) for each participant with two or more time points. The ratio of the mean of the slopes to the standard deviation of slopes was calculated as the Mean-to-Standard Deviation Ratio (MSDR), which provided a standardized measure of signal-to-noise ratio for each feature in detecting progression. A one-sample Wilcoxon signed-rank test was performed on the distribution of individual slopes to determine whether the median progression rate significantly deviated from zero. To compare longitudinal sensitivity across measures, one-sample Wilcoxon signed-rank tests against zero (with FDR-corrected p-values) were applied to slope distributions for the ankle composite, BARS total, and BARS gait, with each participant's slopes computed over the matched time points at which both the sensor composite and clinical scores were available, so that the rates of change were estimated over an identical time interval.

1 Esliger DW, Rowlands AV, Hurst TL, Catt M, Murray P, Eston RG. Validation of the GENEA accelerometer. *Med Sci Sports Exerc* 2011; **43**: 1085–93.

2 Gupta AS, Luddy AC, Khan NC, Reiling S, Thornton JK. Real-life wrist movement patterns capture motor impairment in individuals with ataxia-telangiectasia. *Cerebellum* 2023; **22**: 261–71.

3 Benjamini Y, Hochberg Y. Controlling the false discovery rate: A practical and powerful approach to multiple testing. *J R Stat Soc Series B Stat Methodol* 1995; **57**: 289–300.

4 Mann HB, Whitney DR. On a Test of Whether one of Two Random Variables is Stochastically Larger than the Other. *Ann Math Stat* 1947; **18**: 50–60.

5 Shrout PE, Fleiss JL. Intraclass correlations: Uses in assessing rater reliability. *Psychol Bull* 1979; **86**: 420–8.

6 Koo TK, Li MY. A guideline of selecting and reporting intraclass correlation coefficients for reliability research. *J Chiropr Med* 2016; **15**: 155–63.
